# Genetic markers of enhanced functional antibody responses to COVID-19 vaccination

**DOI:** 10.1101/2025.01.20.25320822

**Authors:** Ruth A Purcell, L Carissa Aurelia, Lilith F Allen, Katherine A Bond, Deborah A Williamson, Janine M Trevillyan, Jason A Trubiano, Bruce D Wines, P Mark Hogarth, Jennifer A Juno, Adam K Wheatley, Thi HO Nguyen, Kanta Subbarao, Katherine Kedzierska, Stephen J Kent, Siddhartha Mahanty, Kevin John Selva, Amy W Chung

## Abstract

**Introduction:** Substantial population-level variation in vaccine-specific antibody responses has been observed following global coronavirus disease 2019 (COVID-19) vaccination efforts. Beyond the influence of clinical and demographic features, immunogenetic variation is suggested to underlie divergent serological responses following COVID-19 vaccination of distinct populations.

**Methods:** Immunoglobulin G1 (IgG1) allotypic markers (G1m) for 121 COVID-19 vaccinated healthy adults were genotyped via Sanger sequencing. Vaccine-specific IgG and Fc gamma receptor (FcγR) engagement were characterised via bead-based multiplex array.

**Results:** Following two COVID-19 vaccine doses, G1m1,17^+/+^ compared to G1m-1,3^+/+^ vaccinees had increased IgG and FcγR engagement specific for the antigenically conserved SARS-CoV-2 Spike 2 (S2) domain. IgG targeting antigenically novel SARS-CoV-2 receptor binding domain (RBD) trended higher in G1m1,17^+/+^ vaccinees, facilitating increased RBD-specific FcγR2a-R131 and FcγR2b binding.

**Conclusion:** Primary COVID-19 vaccination induced increased S2-specific IgG in G1m1,17^+/+^ vaccinees, potentially facilitating enhanced anti-viral FcγR activation. Validation in larger cohorts may inform optimisation of next-generation vaccination strategies.

## Introduction

The immunogenicity of vaccines against coronavirus disease 2019 (COVID-19) is highly heterogenous within and between diverse populations, regardless of vaccine platform [1-4]. Clinical and demographic features, such as chronic comorbidities [3] and older age [4], have been associated with impaired antibody responses following COVID-19 vaccination. However, the potential influence of immunogenetics upon vaccine immunogenicity remains underexplored [5, 6]. Single nucleotide polymorphisms within Immunoglobulin (Ig) heavy chain constant regions, termed IgG allotypes, have previously been associated with modulation of antigen-specific antibody responses following infection with or vaccination against a range of viral, bacterial, and parasitic pathogens [6-12]. Given the geographic clustering of allotypes resulting from a Mendelian inheritance pattern, these genetic markers may inform population-specific vaccination strategies [6].

Beyond neutralising antibody titres—the best described correlate of protection against COVID-19 [13]—other humoral immune responses, including non-neutralising antibodies show considerable variation between vaccinees [1-4]. In contrast to neutralisation, anti-viral Fc effector functions are more durable and better maintained against viral variants since non-neutralising antibodies targeting antigenically conserved regions facilitate cross-protective FcγR activation [14-16]. As such, understanding potential genetic determinants of improved functional antibody responses may inform optimisation of next-generation vaccination strategies. IgG allotypes are proposed to influence IgG1-4 subclass distribution such that the balance of cytophilic (IgG1 and IgG3) and non-inflammatory (IgG2 and IgG4) subclasses is altered [17]. Consequently, increased proportions of cytophilic IgG have the potential to enhance Fc gamma receptor (FcγR) binding and downstream Fc effector functions induced by non-neutralising antibodies [18]. Furthermore, amino acid variation within the antibody Fc region has been reported to influence IgG affinity for FcγRs [19, 20].

Here, within a cohort of COVID-19 vaccinated healthy adults who received primary mRNA-based BNT162b2 or adenoviral vector-based AZD1222 plus mRNA booster, we assessed the influence of Immunoglobulin Heavy Constant Gamma 1 (*IGHG1*) genetic variation upon severe acute respiratory syndrome coronavirus 2 (SARS-CoV-2)-specific IgG levels and FcγR engagement. G1m1,17/G1m1,17 homozygosity was associated with increased IgG and correspondingly increased antibody-mediated FcγR binding specific for the antigenically conserved spike 2 (S2) domain. G1m1,17-associated increases in functional antibody responses against antigenically novel epitopes were less pronounced.

## Methods

### Study participants and sample collection

SARS-CoV-2 vaccine plasma, granulocyte, and PBMC samples were collected from 121 vaccinees 1 month post-second dose BNT162b2 (Pfizer-BioNTech) or AZD1222 (AstraZeneca) vaccination and 1 month post-mRNA booster (BNT162b2 or mRNA-1273 (Moderna)), as previously described [14, 21-23]. Vaccinee cohort demographics are described in Table 1. Study protocols were approved by the University of Melbourne (#2056689, #20734, #21560, #21626, and #13344), Austin Health (#HREC/73256/Austin-2021), and Melbourne Health (HREC/63096/MH-2020 and HREC/68355/MH-2020).

**Table 1.**
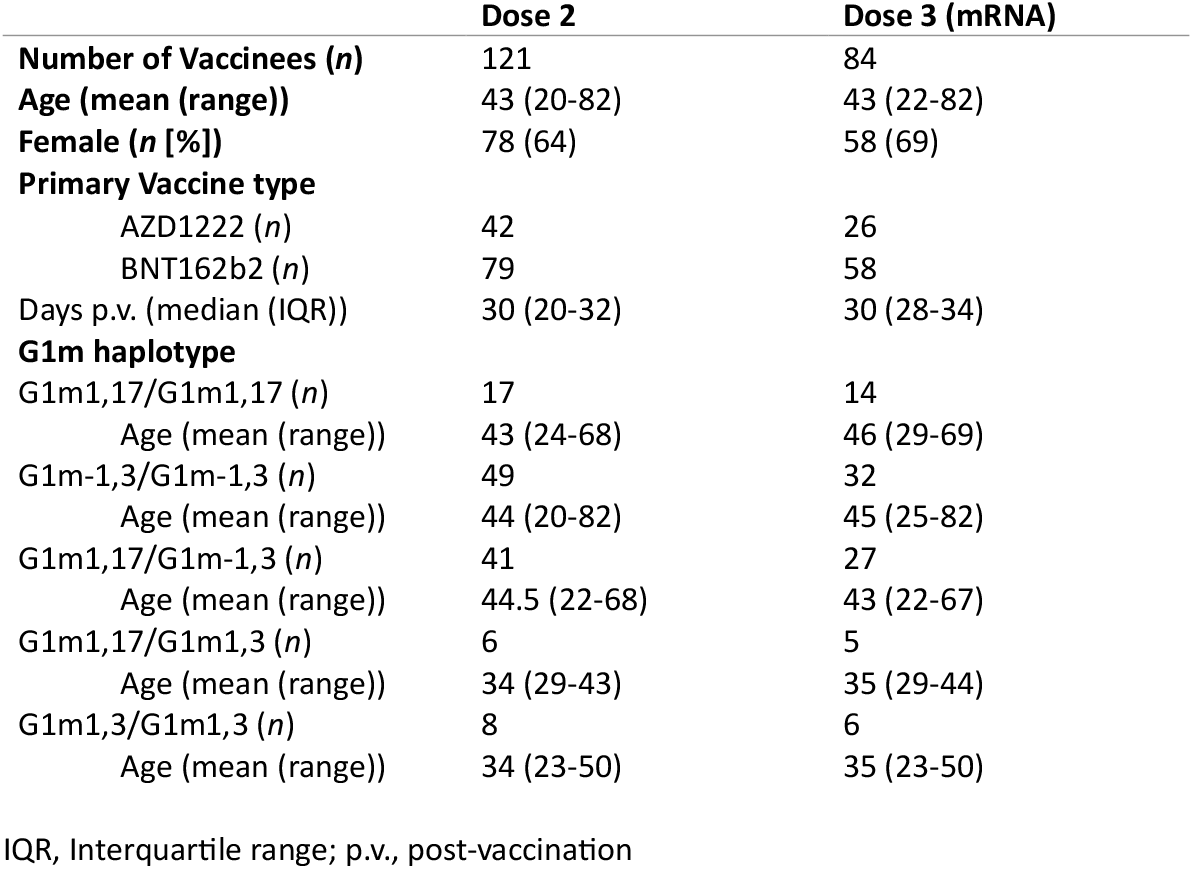
Cohort demographics.

|  | **Dose 2** | **Dose 3 (mRNA)** |
| --- | --- | --- |
| **Number of Vaccinees (n)** | 121 | 84 |
| **Age (mean (range))** | 43 (20-82) | 43 (22-82) |
| **Female (n [%])** | 78 (64) | 58 (69) |
| **Primary Vaccine type** |  |  |
| AZD1222 (n) | 42 | 26 |
| BNT162b2 (n) | 79 | 58 |
| Days p.v. (median (IQR)) | 30 (20-32) | 30 (28-34) |
| **G1m haplotype** |  |  |
| G1m1,17/G1m1,17 (n) | 17 | 14 |
| Age (mean (range)) | 43 (24-68) | 46 (29-69) |
| G1m-1,3/G1m-1,3 (n) | 49 | 32 |
| Age (mean (range)) | 44 (20-82) | 45 (25-82) |
| G1m1,17/G1m-1,3 (n) | 41 | 27 |
| Age (mean (range)) | 44.5 (22-68) | 43 (22-67) |
| G1m1,17/G1m1,3 (n) | 6 | 5 |
| Age (mean (range)) | 34 (29-43) | 35 (29-44) |
| G1m1,3/G1m1,3 (n) | 8 | 6 |
| Age (mean (range)) | 34 (23-50) | 35 (23-50) |

IQR, Interquartile range; p.v., post-vaccination

### DNA extraction, Polymerase chain reaction (PCR), and Sequencing

G1m1/G1m-1 and G1m3/G1m17 typing of study participants was performed via PCR and Sanger sequencing, as previously described [21, 24]. Briefly, genomic DNA was extracted from granulocytes or PBMCs using the QIAamp DNA Blood Mini Kit (Qiagen GmbH, Hilden, Germany; 51104) according to the manufacturer’s instructions. Amplification of the human C_H_1 and C_H_3 domains of *IGHG1* was performed using the AccuPrime *Taq* DNA Polymerase, High Fidelity system (Thermo Fisher Scientific, 12346094). Dual direction sequencing of PCR products was performed by the Australian Genome Research Facility (AGRF, Melbourne, Australia). Geneious Prime version 2023.2.1 was used for sequence analysis and genotypes were manually called.

### Bead-based multiplex assay

To characterise antigen-specific plasma antibody responses, we utilized a customised SARS-CoV-2 antigen multiplex bead array, validated as previously described [14, 21-23]. Briefly, plasma diluted in PBS at an appropriate single concentration (1:100 for dose 2 AZD1222 vaccinee plasma, and 1:800 for dose 2 BNT162b2 vaccinee plasma and all mRNA booster vaccinee plasma) were incubated with antigen-coupled beads (Supplementary Table 1) and the levels of total IgG, IgG1-4 or soluble FcγR dimer engagement (Supplementary Table 2) were assessed via FLEXMAP 3D (Luminex, Austin, TX, USA). Optimal plasma dilutions were determined by calculating the average EC_50_ response specific to each vaccine regimen timepoint and selected for suitability across detectors (Supplementary Figure 1). Anti-IgG and anti-IgG1 antibody detector clones were selected following demonstration of equivalent binding to the different IgG1 allotypes of interest [21]. Detailed methods are described in Supplementary materials.

### Statistical analysis

Prism GraphPad version 10.4.0 (GraphPad Software, San Diego, CA, USA) was used to develop graphs and perform the statistical analyses described in the figure legends.

## Results

### G1m1,17 haplotype associates with increased IgG against SARS-CoV-2 spike 2 domain

We identified five *IGHG1* haplotypes at expected frequencies within our COVID-19 vaccinated cohort (Table 1). Total IgG and IgG1-4 subclass levels against whole SARS-CoV-2 spike trimer (ST) as well as spike 2 domain (S2), spike 1 domain (S1), and receptor binding domain (RBD)— distinct subunits of ST—were then assessed. Importantly, the degree of antigenic conservation with seasonal coronaviruses varies between ST subunits, with S2 being most conserved and RBD being most novel [25]. Following two COVID-19 vaccine doses, G1m1,17/G1m1,17 vaccinees, as compared to G1m-1,3/G1m-1,3 vaccinees, generated increased total IgG against SARS-CoV-2 S2 and ST (Figure 1a-b) as well as increased S2-specific IgG1 (Figure 1e). Although not statistically significant, S1- and RBD-specific total IgG as well as ST-, S1- and RBD-specific IgG1 levels trended higher in G1m1,17/G1m1,17 vaccinees (Figure 1c-d, 1f-h). These data suggest that the influence of IgG1 allotypes upon COVID-19 vaccine responses is antigen-dependent, with the effect being most pronounced in the context of IgG targeting antigenically conserved epitopes.

**Figure 1.**
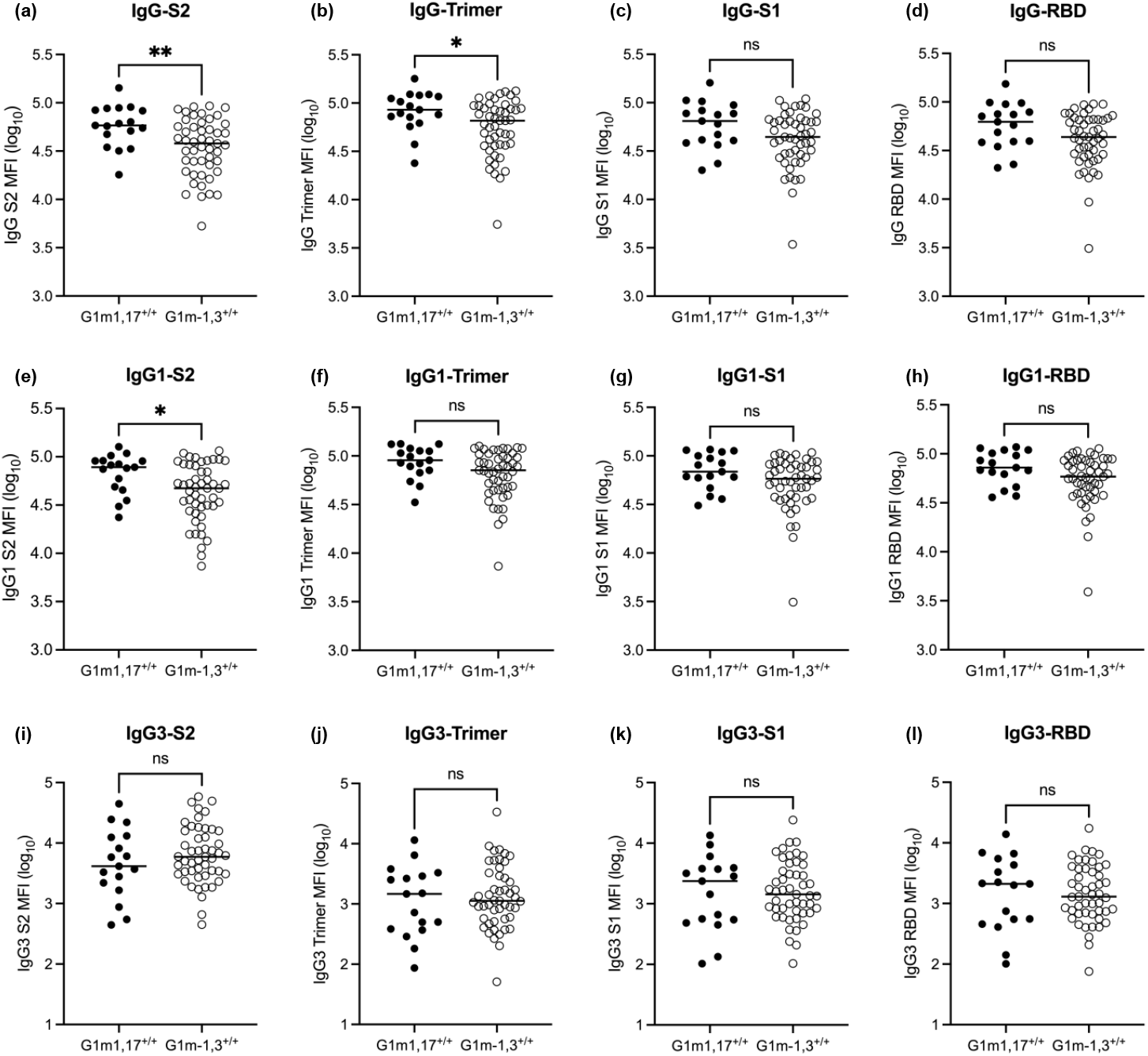
Association of *IGHG1* haplotypes with SARS-CoV-2-specific antibody levels following two COVID-19 vaccine doses. **(a)** S2-, **(b)** Trimer-, **(c)** S1-, and **(d)** RBD-specific IgG levels. **(e)** S2-, **(f)** Trimer-, **(g)** S1-, and **(h)** RBD-specific IgG1 levels. **(i)** S2-, **(j)** Trimer-, **(k)** S1-, and **(l)** RBD-specific IgG3 levels. Mann-Whitney *U*-tests performed between G1m1,17/G1m1,17 and G1m-1,3/G1m-1,3 vaccinees for each antibody-antigen feature. *P* < 0.01 (^**^); *P* < 0.05 (^*^); non-significant (ns).

Upon multiple comparisons between all five *IGHG1* haplotypes, we observed an intermediate anti-S2 IgG response for heterozygotes, albeit with reduced statistical power comparing all haplotypes (Supplementary Figure 2). Following a third COVID-19 vaccine dose, the influence of G1m1,17 homozygosity upon elevated S2-specific IgG was diminished (Supplementary Figure 3).

Using the anti-IgG3 detector clone MTG34, no association between *IGHG1* haplotype and quantity of SARS-CoV-2-specific IgG3—the second most dominant IgG subclass following two BNT162b2 or AZD1222 doses [26, 27]—was observed (Figure 1i-l). As we were unable to identify an IgG3-specific detection antibody that was able to equivalently bind IgG3 allotypes of interest (Supplementary Figure 4), we were unable to assess the potential influence of *IGHG3* haplotypes upon SARS-CoV-2-specific IgG responses.

### Elevated S2-specific IgG facilitates increased FcγR engagement by G1m1,17 homozygous vaccinees

Of the four IgG subclasses, IgG1 and IgG3 possess the highest affinities for FcγRs [28]. This, coupled with the IgG1-dominant response following two COVID-19 vaccine doses [26, 27], underpinned strong correlation of S2-specific total IgG and IgG1 levels with FcγR engagement (Figure 2a). Consequently, the increased anti-S2 IgG, specifically IgG1, observed in G1m1,17/G1m1,17, as compared to G1m-1,3/G1m-1,3, vaccinees facilitated two to threefold increased FcγR2 and FcγR3 binding (Figure 2b-f), although the statistical significance of this effect appeared to be Fc receptor type- and polymorphism-specific.

**Figure 2.**
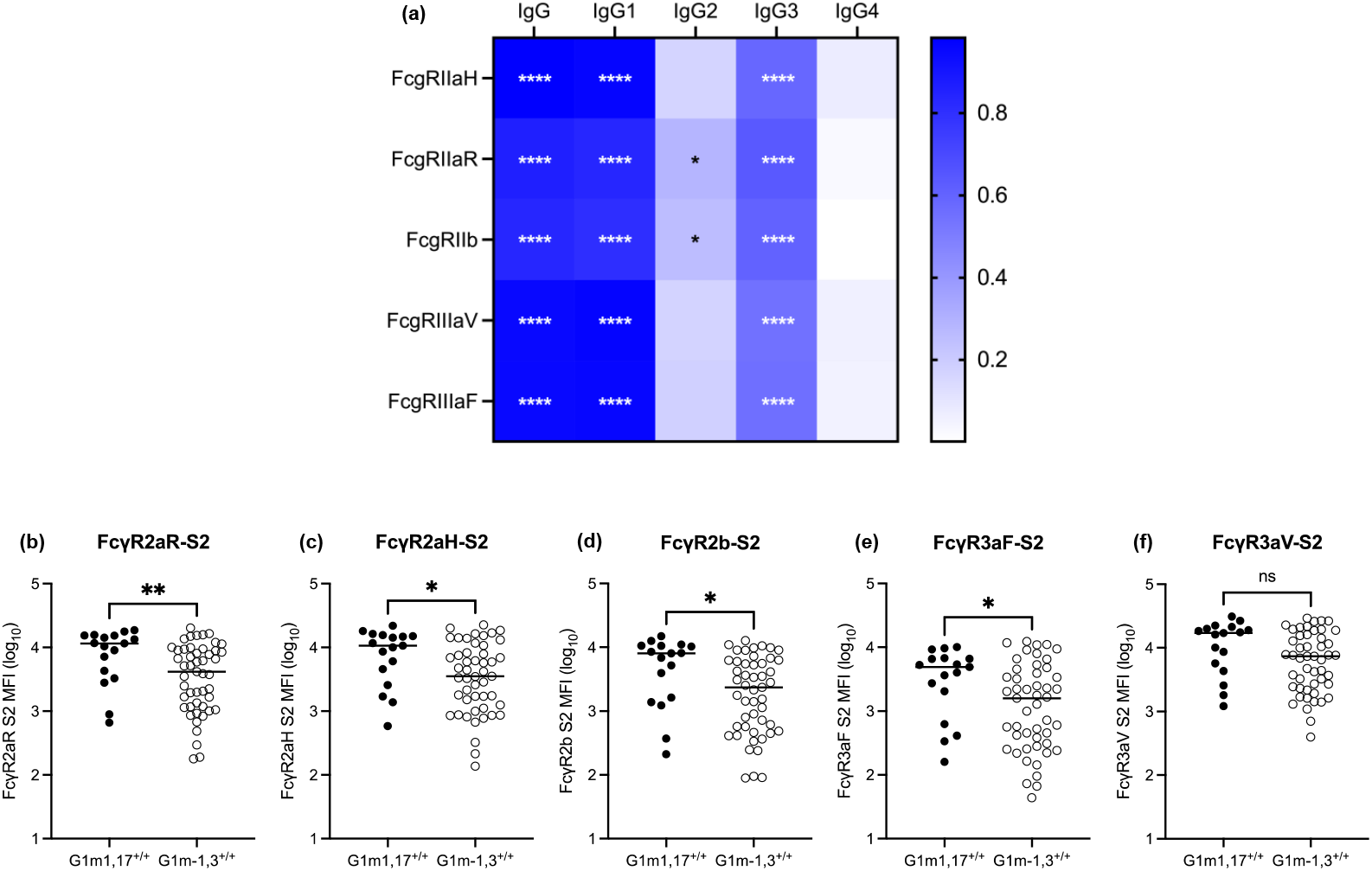
Increased S2-specific IgG levels drive increased S2-specifc FcγR engagement in an FcγR type- and polymorphism-dependent manner. **(a)** Heatmap of spearman correlation coefficients calculated for associations between S2-specifc FcγR engagement and S2-specifc total IgG or IgG subclass levels following two COVID-19 vaccine doses. S2-specifc **(b)** FcγR2aR, **(c)** FcγR2aH, **(d)** FcγR2b, **(e)** FcγR3aF, and **(f)** FcγR3aV engagement following two COVID-19 vaccine doses. Mann-Whitney *U*-tests performed between G1m1,17/G1m1,17 and G1m-1,3/G1m-1,3 vaccinees for each S2-specific FcγR response. *P* < 0.0001 (^****^); *P* < 0.01 (^**^); *P* < 0.05 (^*^); non-significant (ns).

IgG allotypic variation has been associated with differential IgG-FcγR binding and FcγR activation [19, 20]. Previous studies suggest that G1m1,17 IgG1 may bind the FcγR2a-R131 polymorphism with higher affinity than does G1m-1,3 IgG1 [19]. Using biolayer interferometry (BLI), we also observed that the apparent equilibrium dissociation constants (K_D_s) of G1m1,17 IgG1 binding to FcγR2aR and FcγR2b, but not to FcγR2aH, suggested a higher affinity interaction as compared to G1m-1,3 IgG1 (Supplementary Figure 5). For more antigenically the novel SARS-CoV-2 antigens S1 and RBD, as well as ST, significantly increased antibody-mediated FcγR engagement by G1m1,17/G1m1,17 vaccinees was only observed for FcγR2aR and inhibitory FcγR2b, though engagement of other FcγRs trended higher in G1m1,17/G1m1,17 vaccinees (Supplementary Figure 6). Thus, along with differential vaccine-induced IgG subclass titres, *IGHG1* haplotype-specific differences in IgG1-FcγR binding to FcγR polymorphisms may contribute towards variable FcγR engagement.

As predicted by anti-S2 IgG responses, FcγR binding by G1m1,17/G1m-1,3 heterozygous vaccinees represented an intermediate, though not significantly different, response (Supplementary Figure 7). No significant differences in FcγR binding were observed between *IGHG1* haplotypes following dose 3 (Supplementary Figure 3).

## Discussion

In an Australian cohort of COVID-19 naïve healthy adults who received primary BNT162b2 or AZD1222 vaccination, we demonstrated that G1m1,17/G1m1,17 vaccinees generated significantly increased IgG and IgG1 against S2, the most conserved SARS-CoV-2 spike subunit. Responses against more novel antigens such as the RBD trended slightly higher in G1m1,17/G1m1,17 vaccinees. These findings suggest that G1m1,17 haplotype-associated increases in SARS-CoV-2-specific IgG responses may be dependent upon elevated levels of preexisting humoral immune memory to conserved epitopes. We hypothesise that when cross-reactive responses are mounted against an antigen for which prior immune memory exists, for example in this case, against antigenically conserved seasonal coronavirus epitopes, G1m1,17/G1m1,17 vaccinees may have a small but significant advantage in generating a heightened recall antibody response.

However, since trends of elevated S2-specifiic IgG in G1m1,17/G1m1,17 individuals following three COVID-19 vaccines were not statistically significant, G1m haplotypes do not appear to modulate maximal vaccine-specific IgG titres. As such, G1m1,17/G1m1,17 vaccinees may only be advantaged in the setting of boosting a significantly waned memory response (i.e. cross-reactive S2 titres upon primary COVID-19 vaccination) prior to achieving maximal SARS-CoV-2 antigen-specific antibody titres (i.e. following COVID-19 booster vaccination [29]).

Since antigen-specific IgG1 and IgG3 titres are strongly correlated with FcγR2 and FcγR3 binding, we expected that the increased S2-specific total IgG and IgG1 levels observed in G1m1,17/G1m1,17 vaccinees may translate to broadly increased engagement of low-affinity FcγRs. However, the extent of G1m1,17-associated increase in FcγR engagement was dependent upon antigen as well as FcγR type and polymorphism. Significantly increased S2-specific FcγR engagement in G1m1,17/G1m1,17 vaccinees was observed for all tested FcγRs except the higher-affinity polymorphism FcγR3aV. Furthermore, FcγR2aR and FcγR2b were the only FcγRs for which significantly increased ST-, S1-, and RBD-specific FcγR engagement were observed in G1m1,17/G1m1,17 vaccinees. These data suggest that FcγR2 and FcγR3 genetic variation may influence the magnitude of the functional consequence of increased IgG in G1m1,17/G1m1,17 vaccinees.

In agreement with our BLI data, previous data suggest that G1m1,17 IgG1 binds the FcγR2aR polymorphic FcγR2a variant with higher affinity than does G1m-1,3 IgG1 [19]. In prior studies, this effect was not observed for FcγR2aH, FcγR3aV, and FcγR3aF—other low-affinity activating FcγRs—or for high-affinity FcγRI [19]. These data suggest a potential epistatic interaction between the *IGHG1* haplotype G1m1,17 and the lower-affinity FcγR2a polymorphism, FcγR2aR, whereby a combination of these two genotypes may promote increased FcγR2a-driven enhancement of Fc effector functions. However, given that G1m1,17/G1m1,17 vaccinees also demonstrated increased binding to inhibitory receptor FcγR2b, their potential for increased FcγR2aR-mediated effector functions may be overridden. Future studies should confirm the functional consequences of increased FcγR2aR engagement by G1m1,17 IgG1 using cell-based Fc functional assays employing effector cells with known FcγR polymorphisms.

Our study is limited by several technical and cohort-specific restrictions. Given that *IGHG1* and *IGHG3* are in linkage disequilibrium and tend to be inherited as haplotype blocks, IgG1 and IgG3 allotypes have been associated with modulation of antigen-specific levels of the alternate IgG subclass, as well as total antigen-specific IgG [7, 9, 10]. However, as we were unable to identify an anti-IgG3 detector clone that equivalently binds IgG3 allotypes of interest, the influence of IgG3 allotypes, and by extension, IgG1 allotypes, upon IgG3 levels could not be determined reliably. The low frequency of the homozygous G1m1,17/G1m1,17 genotype posed a substantial challenge to recruiting the number of vaccinees required to perform sufficiently powered statistical calculations. This limitation was especially evident at the dose 3 time point for which genotyping identified only 14 G1m1,17/G1m1,17 participants. Given that the Mendelian inheritance pattern of IgG haplotypes facilitates a geographic clustering of genotypes [6], future studies exploring the influence of IgG allotypes upon antibody responses would benefit from multi-centre international cohorts recruited from the specific geographical regions in which the haplotypes of interest are most prevalent.

IgG haplotype-associated differences in antigen-specific Ig titres, Fc functional antibody responses, and protection against disease have been reported for a range of viral, bacterial, and parasitic pathogens [7-12, 17]. While the influence of distinct haplotypes appears to be pathogen-specific [6], the G1m1,17 haplotype has consistently been associated with elevated antibody titres against viral antigens [9, 17]. Here, following two COVID-19 vaccine doses, we demonstrated that G1m1,17 haplotype is associated with a small but significant increase in IgG titres and FcγR engagement, predominantly targeting the antigenically conserved S2 antigen. Larger independent cohort studies are required to determine if these IgG haplotype-associated differences contribute to vaccine efficacy.

## CRediT authorship contribution statement

**Ruth A Purcell:** Conceptualization, Data curation, Formal analysis, Investigation, Methodology, Writing – original draft, Writing – review & editing. **L Carissa Aurelia:** Conceptualization, Data curation, Formal analysis, Investigation, Methodology, Writing – review & editing. **Lilith F Allen:** Investigation. **Katherine A Bond:** Investigation. **Deborah A Williamson:** Investigation, Writing – review & editing. **Janine M Trevillyan:** Investigation, Writing – review & editing. **Jason A Trubiano:** Funding acquisition, Investigation. **Bruce D Wines:** Resources, Writing – review & editing. **P Mark Hogarth:** Resources. **Jennifer A Juno:** Funding acquisition, Investigation, Project administration, Writing – review & editing. **Adam K Wheatley:** Funding acquisition, Investigation, Resources. **Thi HO Nguyen:** Funding acquisition, Investigation, Project administration, Writing – review & editing. **Kanta Subbarao:** Funding acquisition, Investigation, Project administration, Writing – review & editing. **Katherine Kedzierska:** Funding acquisition, Investigation, Project administration, Writing – review & editing. **Stephen J Kent:** Funding acquisition, Investigation, Project administration, Supervision, Writing – review & editing. **Siddhartha Mahanty:** Investigation, Project administration, Writing – review & editing. **Kevin John Selva:** Investigation, Supervision, Writing – review & editing. **Amy W Chung:** Conceptualization, Formal analysis, Funding acquisition, Investigation, Methodology, Project administration, Supervision, Writing – review & editing.

## Acknowledgements

This study was supported by a Medical Research Future Fund (MRFF) GNT #2016062 to JAT, JAJ, AKW, THON, KK, SJK, and AWC, and a National Health and Medical Research Council (NHMRC) Investigator grant #2008092 to AWC. JAJ, AKW, THON, KS, KK, and SJK are also supported by NHMRC Investigator grants. JAJ is also supported by the Charles and Sylvia Viertel Charitable Foundation. The DISCOVER-HCP study was supported by the National Institute of Allergy and Infectious Diseases, National Institutes of Health, Department of Health and Human Services, Centers of Excellence for Influenza Research and Response (CEIRR) grant #HHSN272201400005C to the University of Rochester and a subcontract to KS. This work was made possible through Victorian State Government Operational Infrastructure Support and Australian Government NHMRC IRIISS. We thank P Ramanathan, E Haycroft, T Amarasena, K Wragg, P Konstandopoulos, G Gare, K Field, H Kelly (University of Melbourne), G Gibney, and F James (Austin Health) for their outstanding technical assistance.

## Data availability

The data that support the findings of this study are available from the corresponding author upon reasonable request.

