## Supplementary Tables and Figures for "Genetic markers of enhanced functional antibody responses to COVID-19 vaccination"

**Supplementary Table 1.** Details of ancestral SARS-CoV-2 antigens coupled to magnetic carboxylated beads used in bead-based multiplex assays

| Antigen | μg/1.25×$10^7$ beads | Provider | Catalogue number |
|---|---|---|---|
| Spike Trimer (ST) | 30 | Produced in-house | PMID: 32661393 |
| Spike 2 (S2) | 100 | ACROBiosystems | S2N-C52H5 |
| Spike 2 (S1) | 100 | Sino Biological | 40591-V08H |
| Receptor Binding Domain (RBD) | 100 | Sino Biological | 40592-V08H |

**Supplementary Table 2.** Details of mouse anti-human anti-IgG antibody detectors used in bead-based multiplex assays

| Detector | Clone | Provider | Catalogue number |
|---|---|---|---|
| Total IgG, PE | JDC-10 | Southern Biotech | 9040-09 |
| IgG1, PE | HP6001 | Southern Biotech | 9054-09 |
| IgG2, biotin | HP6200 | MabTech | 3852-6-250 |
| IgG3, biotin | MTG34 | MabTech | 3853-3-250 |
| IgG4, PE | HP6025 | Southern Biotech | 9200-09 |
| FcγR2a-H131 | N/A | Produced in-house | PMID: 27385782 |
| FcγR2a-R131 | N/A | Produced in-house | PMID: 27385782 |
| FcγR2b | N/A | Produced in-house | N/A |
| FcγR3a-V158 | N/A | Produced in-house | PMID: 27385782 |
| FcγR2a-F158 | N/A | Produced in-house | PMID: 27385782 |

PE, R-Phycoerythrin. Responses detected via PE-conjugated antibodies were read directly. Responses detected via biotinylated antibodies were read following an additional incubation with 1 μg $mL^{-1}$ Streptavidin, R-Phycoerythrin Conjugate (SAPE) (Thermo Fisher Scientific, S866).

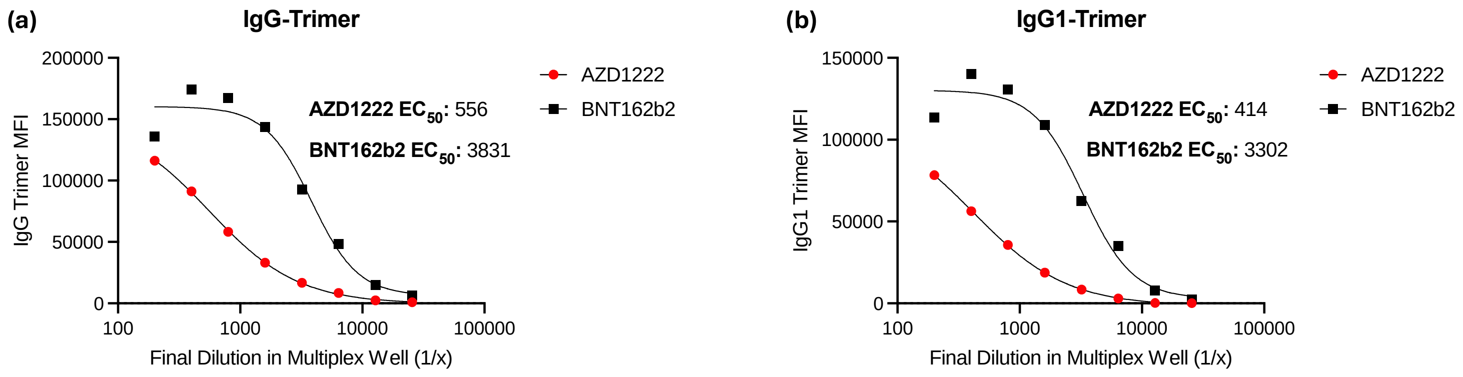


**Supplementary Figure 1.** Titration curves for SARS-CoV-2 Spike antigen array. Titration curves depict **(a)** total IgG and **(b)** IgG1 responses against ancestral Spike Trimer using pooled plasma from representative AZD1222 dose 2 vaccinees (red circles) and BNT162b2 dose 2 vaccinees (black squares). Curves were fitted using a four-parameter nonlinear regression. Median Fluorescence Intensity (MFI).

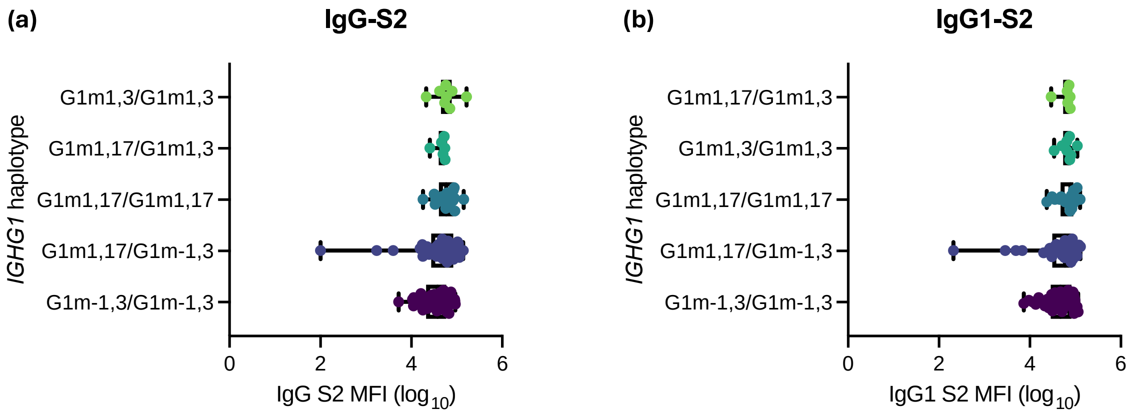


**Supplementary Figure 2.** Association of five dominant *IGHG1* haplotypes with SARS-CoV-2 S2-specific **(a)** IgG levels, and **(b)** IgG1 levels. No significant differences in S2-specific IgG or IgG1 levels between any *IGHG1* haplotypes were observed following multiple comparisons performed via Kruskal-Wallis test. Median Fluorescence Intensity (MFI).

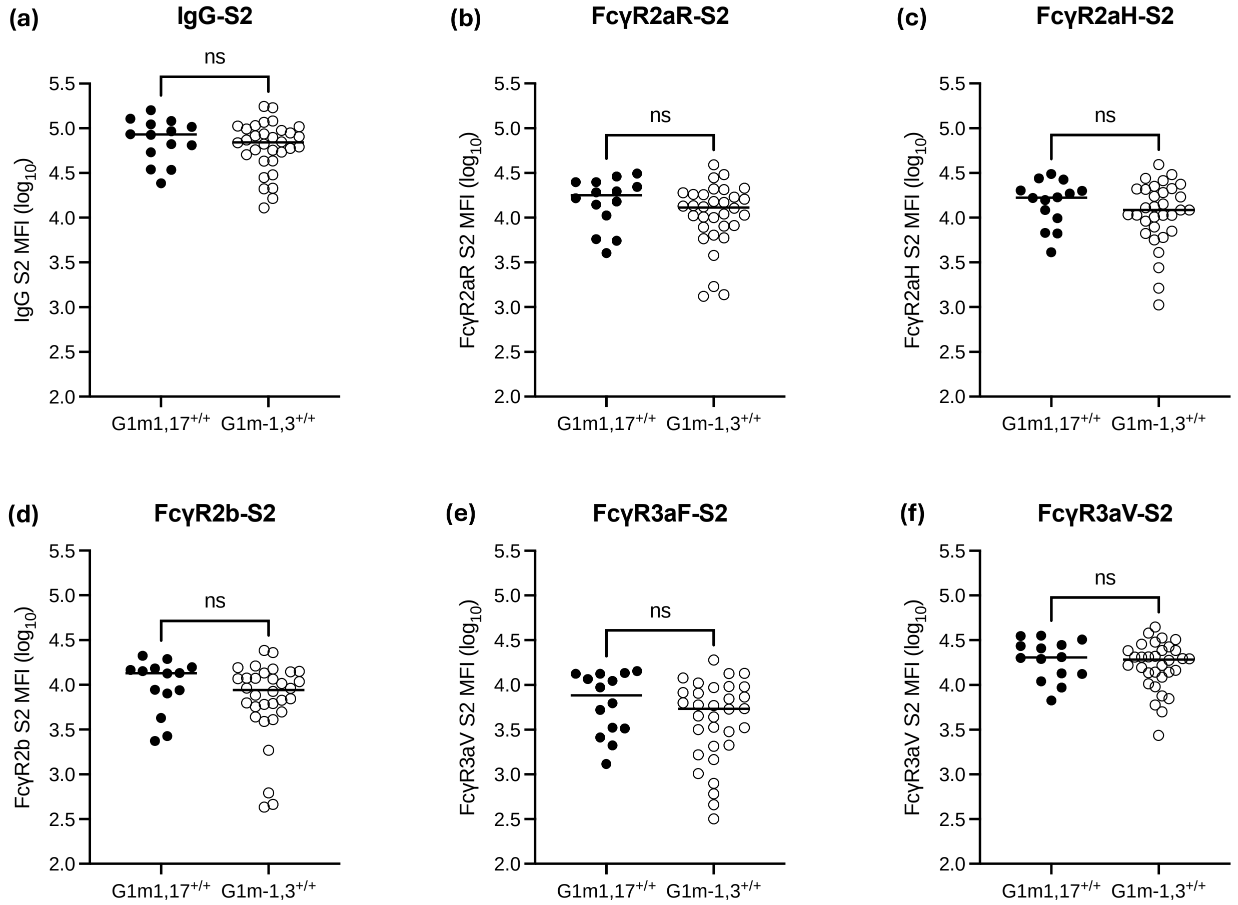


**Supplementary Figure 3.** Association of *IGHG1* haplotypes with SARS-CoV-2 S2-specific **(a)** IgG levels, **(b)** FcγR2aR engagement, **(c)** FcγR2aH engagement, **(d)** FcγR2b engagement, **(e)** FcγR3aF engagement, and **(f)** FcγR3aV engagement following three COVID-19 vaccine doses**.** Mann-Whitney *U*-tests performed between G1m1,17/G1m1,17 ($n$ = 14) and G1m-1,3/G1m-1,3 ($n$ = 32) vaccinees for each antibody- or FcγR- antigen feature. non-significant (ns). Median Fluorescence Intensity (MFI).

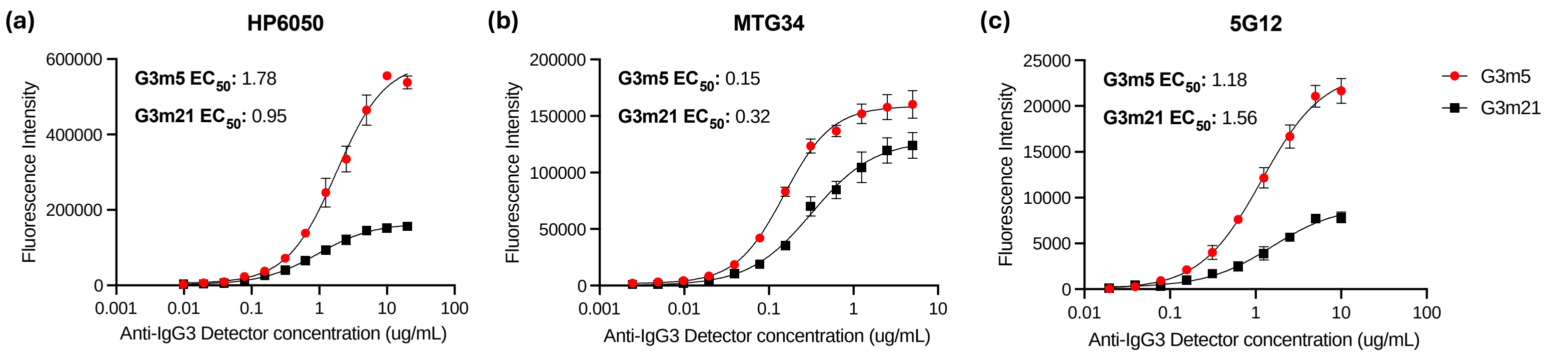


**Supplementary Figure 4.** Neither HP6050, MTG34, nor 5G12 anti-IgG3 detection antibody clones bind G3m5 and G3m21 allotype variants equivalently. $EC_{50}$s determined for **(a)** HP6050, **(b)** MTG34, and **(c)** 5G12 anti-IgG3 clones binding G3m5 and G3m21 allotype variants of myeloma IgG3 purified from human plasma (G3m5: red circles; G3m21: black squares). Measurements were performed in triplicate and mean values ± SEM are indicated. Curves were fitted using a four-parameter nonlinear regression. None of the three tested detectors bound G3m5 and G3m21 variants of IgG3 with equivalent capacity, as determined via comparisons of $EC_{50}$s and maximum fluorescence intensities calculated for G3m5 and G3m21. Clone MTG34 was selected for assessment of IgG3 levels in the COVID-19 vaccination cohort since this detector gave the most similar $EC_{50}$s and maximum fluorescence intensities for G3m5 and G3m21 IgG3 variants.

Methodology for assessment of G3m allotype-specific binding capacity of anti-IgG3 detectors is outlined in Supplementary Methods below.

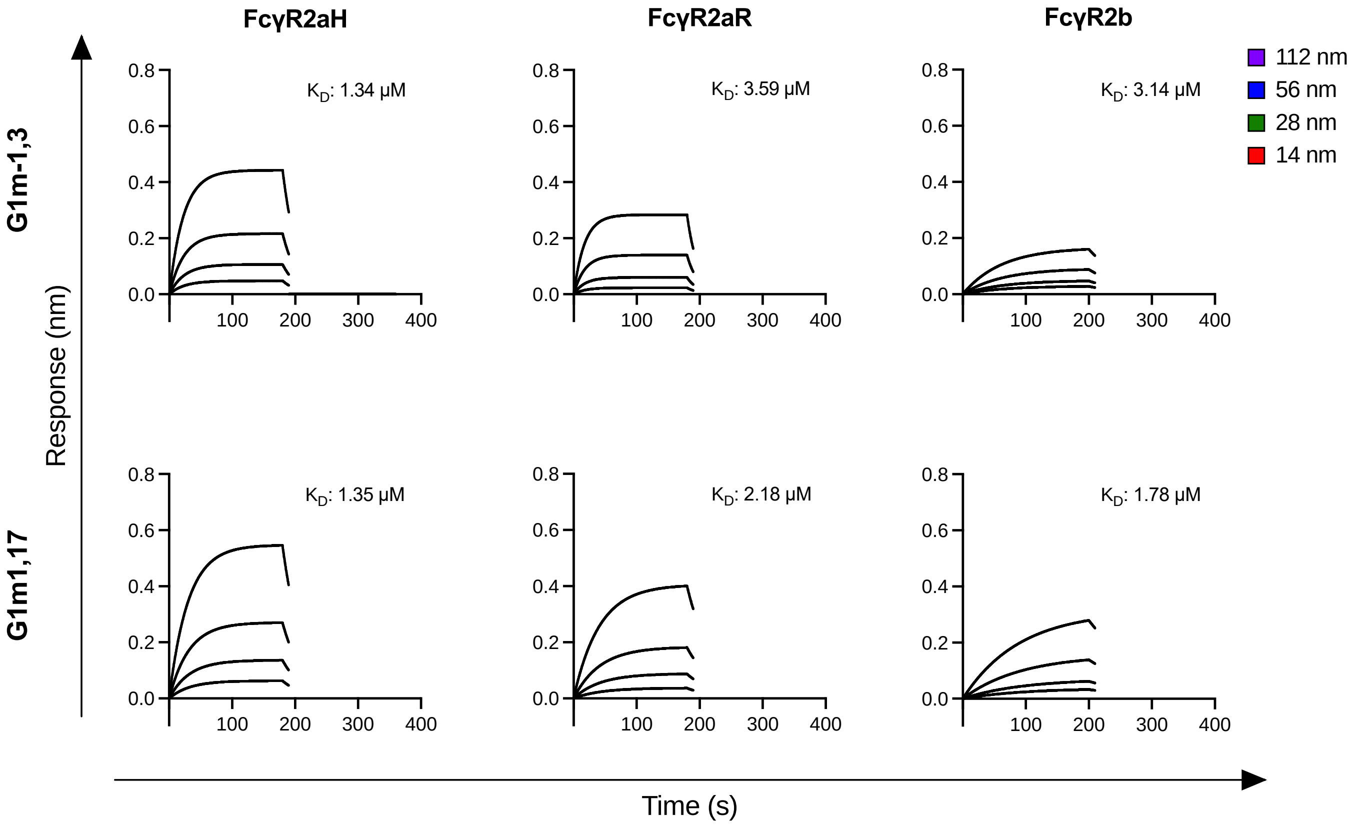


**Supplementary Figure 5.** Increased binding of FcγR2aR and FcγR2b, but not FcγR2aH, by G1m1,17 variant IgG1. Sensograms depict binding of twofold serial dilutions of **(a)** G1m-1,3 allotype IgG1 monoclonal antibody standard and **(b)** G1m1,17 allotype IgG1 monoclonal antibody standard to immobilised FcγR2aH, FcγR2aR, and FcγR2b. Curve fitting was performed using a global fit 1:1 Langmuir binding model. Representative plots of two replicate measurements are shown. Affinity constant/Equilibrium dissociation constant (KD); Micromolar (µM); Nanometre (nm).

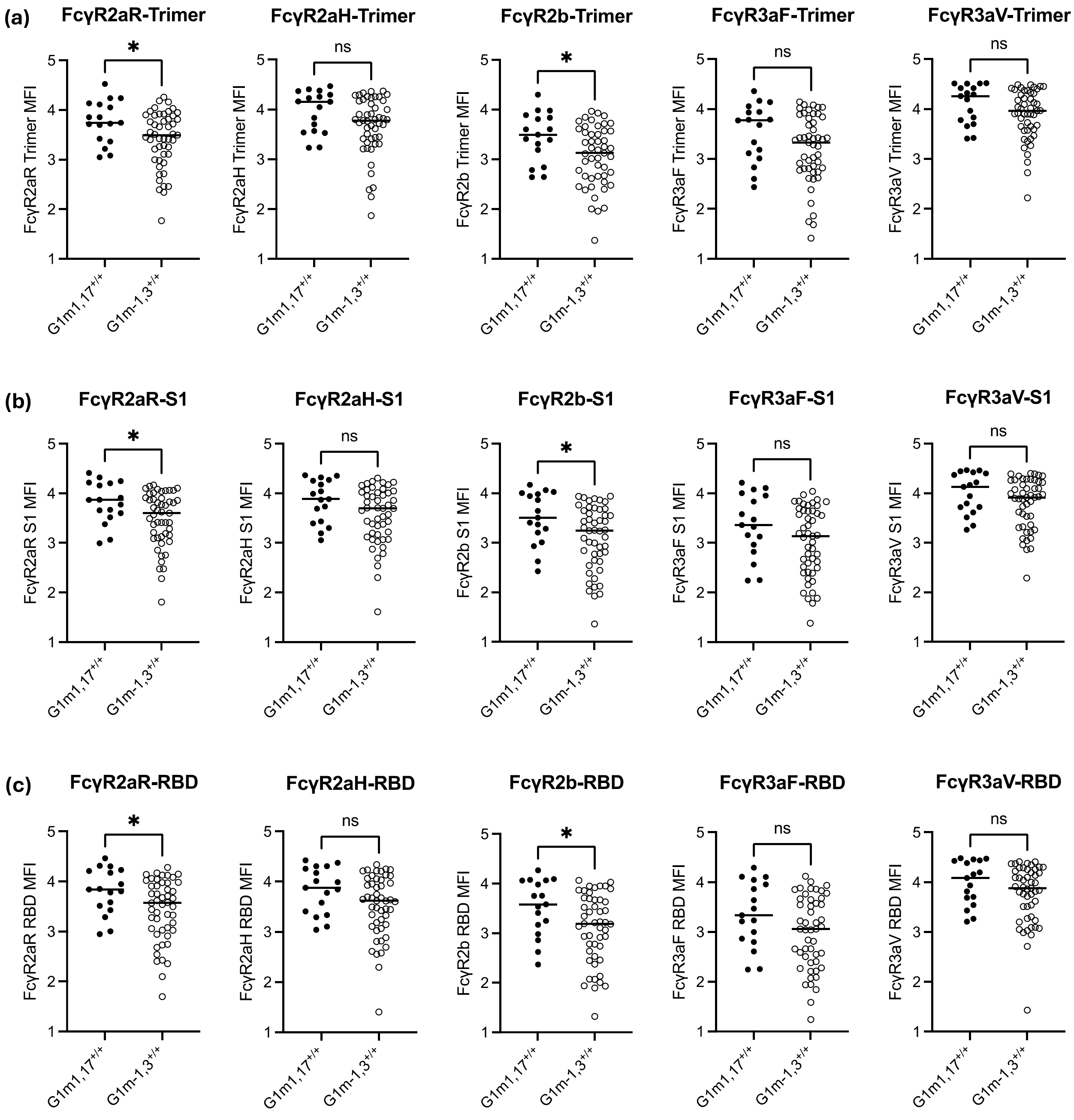


**Supplementary Figure 6.** Association of *IGHG1* haplotypes with SARS-CoV-2 **(a)** Trimer-specific, **(b)** S1-specific, **(c)** RBD-specific FcγR2aR, FcγR2aH, FcγR2b, FcγR3aF, and FcγR3aV engagement following two COVID-19 vaccine doses**.** Mann-Whitney *U*-tests performed between G1m1,17/G1m1,17 ($n$ = 17) and G1m-1,3/G1m-1,3 ($n$ = 49) vaccinees for each antibody- or FcγR- antigen feature. $P < 0.05$ (*); non-significant (ns). Median Fluorescence Intensity (MFI).

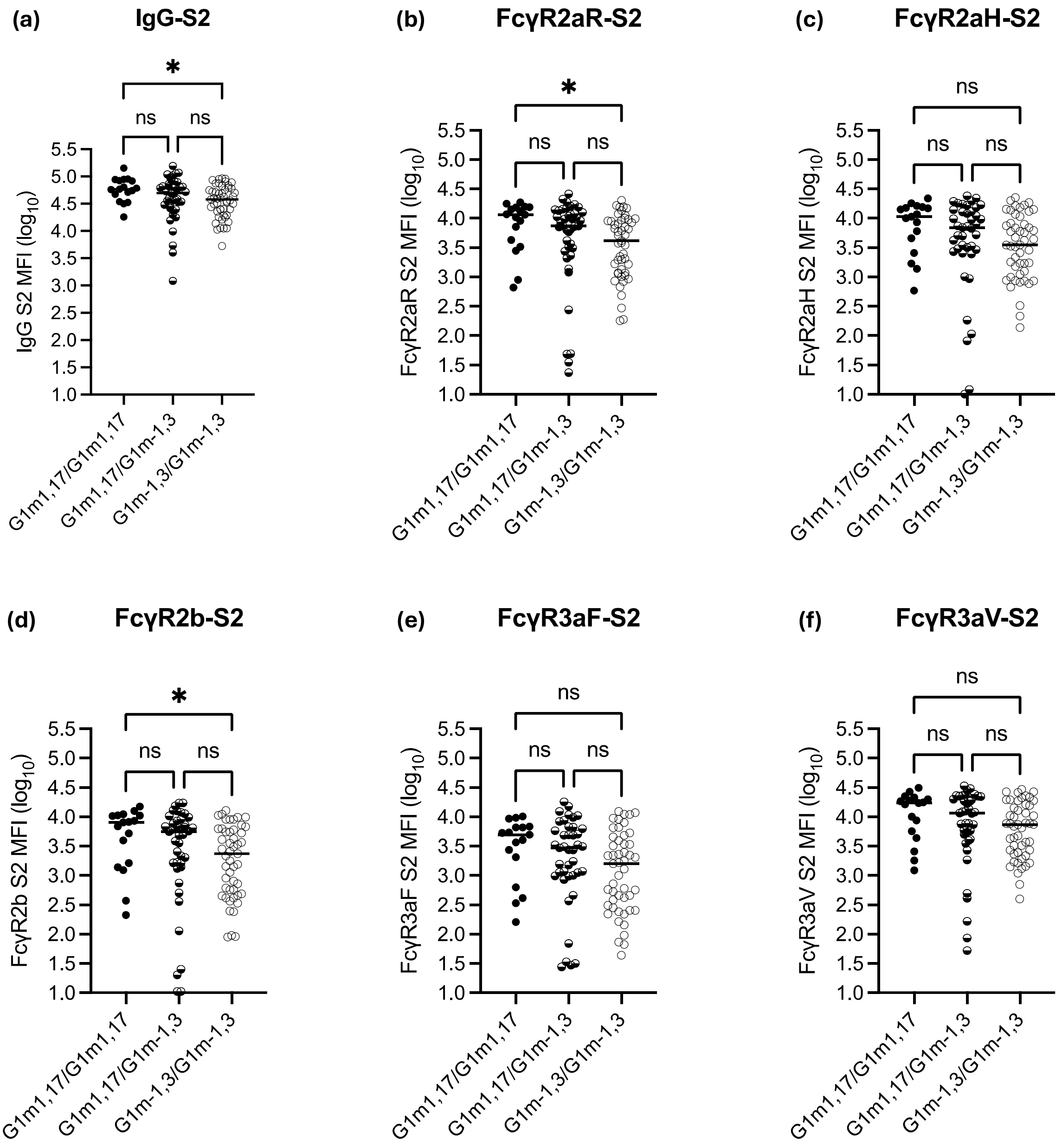


**Supplementary Figure 7.** Association of *IGHG1* haplotypes with SARS-CoV-2 S2-specific **(a)** IgG levels, **(b)** FcγR2aR engagement, **(c)** FcγR2aH engagement, **(d)** FcγR2b engagement, **(e)** FcγR3aF engagement, and **(f)** FcγR3aV engagement following two COVID-19 vaccine doses. Kruskal-Wallis tests performed between G1m1,17/G1m1,17 ($n$ = 17), G1m1,17/G1m-1,3 ($n$ = 41), and G1m-1,3/G1m-1,3 ($n$ = 49) vaccinees for each antibody- or FcγR- antigen feature. $P$ < 0.05 (*); non-significant (ns). Median Fluorescence Intensity (MFI).

**Supplementary Methods**

***Bead-based multiplex assay***

To characterise antigen-specific plasma antibody responses, we utilised a customised SARS-CoV-2 antigen multiplex bead array. Magnetic carboxylated beads (Bio-Rad) were coupled with 100 µg antigen (SARS-CoV-2 Spike 1, Spike 2, or receptor binding domain (Sino Biological, Beijing, China)) or 30 µg Spike Trimer (produced in-house) per $1.25 \times 10^7$ beads. 1000 beads/bead region diluted in PBS containing 0.1% BSA (incubation buffer) were added to each well (total of 25 µL per well) in black, clear-bottom 384-well plates (Greiner Bio-One, Kremsmünster, Austria; 781,906), followed by addition of 25 µL per well of plasma diluted in PBS at an appropriate single concentration, as determined by the average $EC_{50}$ response specific to each vaccine regimen and timepoint, and optimised for suitability across the broadest range of IgG and FcγR detectors (1:100 for dose 2 AZD1222 vaccinee plasma, and 1:800 for dose 2 BNT162b2 vaccinee plasma and all mRNA booster vaccinee plasma) (Supplementary Figure 1). Plates were incubated on a plate shaker overnight at 4 °C before wells were washed with PBS 0.05% Tween-20 (PBST). PE-conjugated mouse anti-human *pan*-IgG (JDC-10 (Southern Biotech, 9040-09)), IgG1 (HP6001 (Southern Biotech, 9054-09)), IgG2 (HP6200 (MabTech, 3852-6-250)), IgG3 (MTG34 (MabTech, 3853-3-250)), IgG4 (HP6025 (Southern Biotech, 9200-09)) or biotinylated FcγR dimers (FcγR2aH, FcγR2aR, FcγRb, FcγR3aV, FcγR3aF (recombinantly expressed in-house)) were diluted to 1.3 µg $mL^{-1}$ and added at 25 µL volume per well before incubation for 2 h at room temperature on a plate shaker. For biotinylated FcγRs, plates were subjected to an additional PBST wash, and then incubated with 1 µg $mL^{-1}$ Streptavidin, R-Phycoerythrin Conjugate (SAPE) (Thermo Fisher Scientific, S866) at room temperature for 2 h. Plates were then washed with PBST and beads were resuspended in 50 µL of sheath fluid per well. The level of PE signal associated with each bead region in each well, reported as median fluorescence intensity (MFI), was determined by a FLEXMAP 3D Luminex instrument system (Luminex, Austin, TX, USA). Each sample was run in duplicate.

***Bio-layer interferometry (BLI)***

Binding kinetics BLI of monoclonal antibody (mAb) allotype standards to human FcγRs were performed on the Octet RED96e instrument (Sartorius, Dandenong South, VIC, Australia) in black 96-well plates (Greiner Bio-One) at 30°C agitated at 1000 RPM. HBS-EP buffer (containing 0.01 M HEPES, 0.15 M NaCl, 3 mM EDTA, 0.005% v/v Surfactant P20, and 0.005% Tween 20; pH 7.4) (GE Healthcare) was used as the kinetic buffer. Biotinylated FcγRIIa, FcγRIIb or FcγRIIIa (5µg/mL) were loaded onto Streptavidin (SA) Biosensors (Sartorius) hydrated in HBS-EP buffer until a threshold of 2 nm was reached, followed by equilibration in buffer only wells for 120 s. The sensors were loaded into a two-fold dilution series of IgG1 allotype mAb standards ((G1m-1,3 kappa, Clone: AbD18705_hIgG1 (Bio-Rad, Gladesville, Australia; HCA192); G1m1,17 kappa, Clone: AbD18705il (Bio-Rad, HCA319)) from 14-112 nM for 180 s to measure association. Dissociation was measured by submerging the sensors into buffer only wells for 180 s. Curve fitting was performed using a global fit 1:1 Langmuir binding model using Octet Data Analysis software v12.0.2.3 (FortéBio), and baseline drift was corrected by reference subtracting the shift of an FcγR-loaded sensor immersed in kinetic buffer only. Mean kinetic constant values from 2 independent experiments were determined, with all binding curves matching the theoretical fit with an $r^2$ value of more than 0.99.

***Enzyme-linked immunosorbent assay (ELISA)***

Nunc MaxiSorp flat-bottom 96-well plates (Thermo Fisher Scientific, Scoresby, Australia; 44-2404) were coated overnight at 4 °C with 50 ng per well of respective purified G3m5 or G3m21

IgG3 allotype standard (G3m5: IgG3, kappa, Human Myeloma, Allotype G3m(b) (US Biological, Salem, MA, USA; I1904-85B3; G3m21: IgG3 lambda, Human Myeloma, Allotype G3m(g) (I1904-85B4)), washed with PBST, then blocked with PBS containing 0.1% BSA (Sigma-Aldrich, St. Louis, MO, USA; A7906-100G) and 0.05% Tween 20 (Sigma-Aldrich, P1379-500ML), and again washed with PBST. Twofold dilutions of anti-IgG3 detection antibodies (Mouse Anti-Human IgG3 Hinge-PE (HP6050) (Southern Biotech, Birmingham, AL, USA; 9210-09); 5G12 (Bio-Rad Laboratories, South Granville, NSW, Australia; 5247-9850); MTG34 (MabTech, Cincinnati, OH, USA; 3853-6-250) were prepared in blocking solution (PBST containing 0.1% BSA), beginning at 5 µg $mL^{-1}$ for MTG34, beginning at 10 µg $mL^{-1}$ for 5G12, and beginning at 20 µg $mL^{-1}$ for HP6050. Anti-human IgG3 antibody detection reagent dilutions were incubated with IgG3 allotype standards coated on the ELISA plate for 2 h at room temperature. ELISAs performed with biotinylated MTG34 and 5G12 were washed five times with PBST then incubated for a further 2 h at room temperature with 1 µg $mL^{-1}$ Streptavidin, R-Phycoerythrin Conjugate (SAPE) (Thermo Fisher Scientific, S866), followed by a final 5 washes with PBST. ELISAs performed with R-Phycoerythrin (PE)-conjugated 4E3 and HP6050 were directly read following a final 5 washes with PBST. Fluorescence intensity was read on a CLARIOstar *Plus* (BMG Labtech, Mornington, Australia) microplate reader with excitation wavelength of 488-15 nm and emission of 576-20 nm using the Enhanced Dynamic Range (EDR) function, selected based on the excitation (496 nm and 565 nm) and emission (578 nm) maxima of PE. Technical replicates were performed in triplicate.
